# Association Between Continuous Glucose Monitoring–Derived Time in Range and Diabetic Kidney Disease: A Systematic Review and Meta-Analysis of current evidence

**DOI:** 10.64898/2026.09.21.26363573

**Authors:** Siddharth Manish Lodha, Sumit Kumar, Shubho Acharya, Parth Dhamelia, Sandeep Garg

**Author notes:** Joint first authors (Siddharth Manish Lodha, Sumit Kumar). Corresponding author: Dr Sandeep Garg, Director Professor, Department of Internal Medicine, Maulana Azad Medical College, New Delhi – 110002, India. email id.

## Abstract

**Backgroun**d**:** Continuous glucose monitoring–derived time in range (TIR; glucose 70-180 mg/dL) has emerged as a complementary glycemic metric, but its association with DKD (Diabetic Kidney Disease) has not been systematically quantified. We aimed to synthesize the evidence on the association between CGM-derived TIR and DKD in adults with type 1 or type 2 diabetes by conducting a systematic review and meta-analysis of the current evidence.

**Methods:** This systematic review and meta-analysis adhered to PRISMA guidelines. PubMed, Embase, Cochrane CENTRAL, and Scopus were searched through March, 2026. Eligible studies were studies in adults with type 1 and type 2 diabetes that reported an association between CGM-derived TIR and any DKD outcome (albuminuria, eGFR decline, or composite nephropathy) with a quantifiable per-unit effect estimate. Random-effects meta-analysis (REML with Knapp-Hartung adjustment) pooled odds ratios per 10% TIR increment. Risk of bias was assessed using ROBINS-E and certainty of evidence using the GRADE framework. PROSPERO: CRD420261377763.

**Results:** Eleven studies (n = 7,182; seven T2D, four T1D; ten cross-sectional, one retrospective cohort) were included. The pooled OR per 10% TIR was 0.89 (95% CI 0.84-0.95; P = 0.002; I² = 69.6%), and every leave-one-out iteration preserved significance (OR range 0.88-0.91; all P ≤ 0.005). In the four studies adjusting for HbA1c, the association persisted (OR 0.91, 0.86-0.97; P = 0.015) with no detectable residual heterogeneity (I² = 0%), indicating the signal is not fully attributable to average glycemia.

**Conclusions:** Higher CGM-derived TIR may be associated with lower odds of DKD (OR 0.89 per 10% increment). The HbA1c-adjusted subgroup (k = 4; I² = 0%) provided the most internally consistent signal, though its small size limits inference. Prospective studies with standardized CGM protocols and pre-specified hard kidney endpoints are needed to establish causality.

## INTRODUCTION

Diabetic kidney disease (DKD), characterized by persistent albuminuria and progressive decline in glomerular filtration rate, develops in 20–40% of individuals with type 1 (T1D) or type 2 (T2D) diabetes and is the leading cause of chronic kidney disease (CKD) and end-stage renal disease worldwide [1–4]. Hemoglobin A1c (HbA1c) has been the cornerstone metric for guiding glycemic management to prevent microvascular complications since the DCCT and UKPDS [5–7]. However, HbA1c represents a weighted average that does not distinguish patients with stable glucose profiles from those with wide swings between hypo- and hyperglycemia, the ‘fallacy of average’ [8], is confounded in anemia, iron deficiency, hemoglobinopathies and CKD itself [9], and does not capture the multiple facets of dysglycemia (postprandial excursions, glycemic variability, hypoglycemia) relevant to microvascular risk [10].

Continuous glucose monitoring (CGM)-derived time in range (TIR; the proportion of time with glucose 70–180 mg/dL [3.9–10.0 mmol/L]) has emerged as a complementary glycemic metric endorsed by international consensus [11,12]. In its 2026 Standards of Care, the ADA expanded CGM recommendations to include adults with type 2 diabetes on non-insulin therapies and CGM initiation at diabetes diagnosis [13], broadening the population in whom CGM-derived metrics including TIR will inform routine clinical care. TIR is a validated surrogate for microvascular risk [14]. TIR is associated with cardiovascular mortality, carotid intima-media thickness, peripheral neuropathy, and retinopathy [15–19], plausibly via oxidative stress and endothelial dysfunction accentuated by glucose oscillations [20], with advanced glycation end-product–mediated injury implicated in DKD pathogenesis [21]. Kidney-specific evidence has been reported [22–26] but varies in CGM protocols (3–28 days), outcome definitions, and HbA1c adjustment. Previous CGM reviews have addressed diabetes complications broadly [27] but none has specifically pooled TIR-DKD estimates. We conducted a systematic review and meta-analysis quantifying this association across T1D and T2D, pre-specifying subgroup and sensitivity analyses, following PRISMA 2020 guidelines [28] with prospective PROSPERO registration.

## METHODS

This review followed PRISMA 2020 guidelines and was prospectively registered with PROSPERO (CRD420261377763); no amendments were made. PubMed, Embase, Cochrane CENTRAL, and Scopus were searched through March 2026 using broad CGM/TIR and diabetes terms without kidney filters [29]; full strategies are in the Supplementary Material. Using the PECO framework, eligible studies enrolled ambulatory adults (≥18 years) with T1D or T2D, measured CGM-derived TIR (70–180 mg/dL), and reported DKD outcomes (albuminuria [UACR ≥30 mg/g, primary], reduced eGFR, CKD progression, or composite nephropathy). Observational designs reporting any association between CGM-derived TIR and DKD were eligible. Non-English publications, pediatric/gestational/ICU populations, SMBG-derived TIR, binary-only comparisons, and duplicate publications were excluded. Two reviewers (S.M.L. and S.K.) independently screened and extracted data, with disagreements resolved by consensus (Figure 1). Extracted data included design, population, sample size, HbA1c, TIR, CGM system/duration, outcome definition, effect estimate with 95% CI, and covariates. Per-1% estimates were converted to per 10% by exponentiation; the most adjusted model was selected. No study authors were contacted. Risk of bias was assessed using ROBINS-E [30] by two independent reviewers across seven domains (Supplementary Table S1). Studies were included in the meta-analysis if they reported an OR, HR, or RR with 95% CI per unit TIR; studies with incompatible effect measures were synthesized narratively. Effect estimates were expressed as OR per 10% TIR on the log scale. One study (Zhang Q 2025 [25]) reported a hazard ratio; the HR was converted to an OR using the Greenland formula [31] with delta-method SE transformation; sensitivity analyses using the unconverted HR and excluding Zhang were also conducted. Random-effects models used REML with the Knapp–Hartung adjustment [32,33]. Heterogeneity was assessed by Cochran’s Q (P<0.10), I², τ², and 95% prediction intervals. Pre-specified subgroups included diabetes type, HbA1c-adjustment status, geographic region, outcome definition, and study design. Univariate meta-regression examined seven moderators (Supplementary Table S4). Sensitivity analyses included leave-one-out, High-RoB exclusion, influence diagnostics, and estimator robustness (Supplementary Table S3). E-values quantified robustness to unmeasured confounding [34]. Publication bias was assessed using Egger’s and Begg’s tests and Duval–Tweedie trim-and-fill. Certainty of evidence was assessed using GRADE, starting at Low for observational studies, with upgrades for plausible confounding direction [35] (Supplementary Table S2). Analyses used R 4.5.3 with metafor 5.0-1 [32]; P < 0.05 was significant except for Q and Egger’s test (P<0.10).

**Fig. 1.**
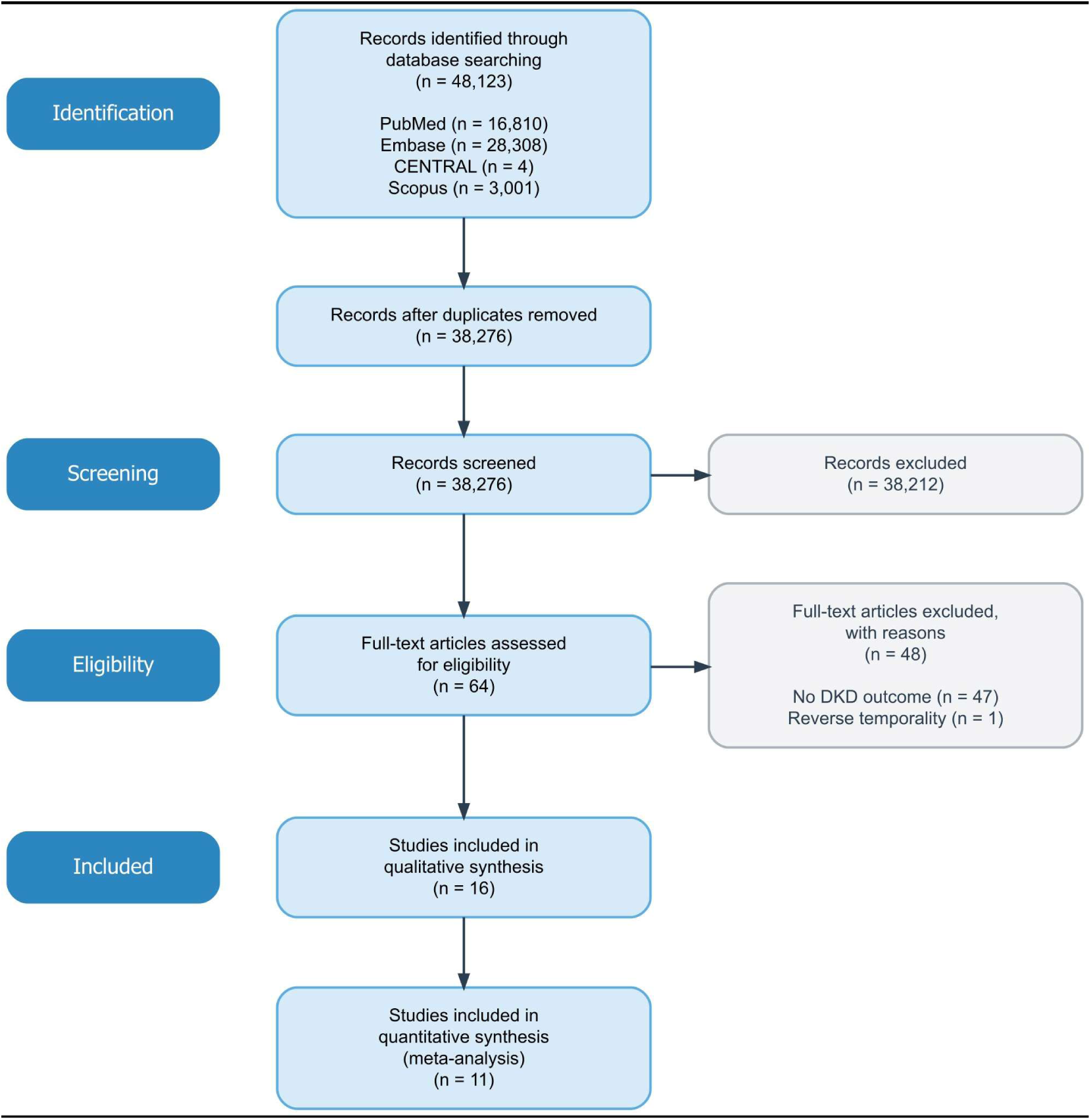
PRISMA 2020 flow diagram presenting the identification, screening, and selection of eligible studies. Reverse temporality denotes a study in which CGM was measured after the kidney outcome was ascertained, precluding causal inference.

## RESULTS

Searches yielded 48,123 records; after deduplication and screening, 64 underwent full-text review and 16 studies met inclusion criteria; 11 (N = 7,182) reported poolable effect estimates and comprised the analytic pool (Figure 1). Five included studies reported TIR–kidney associations but were synthesized narratively owing to incompatible effect measures: unadjusted coefficients only [36], non-standard TIR categories [37], matrix-factorization profiling [38], no direct per-unit TIR–kidney estimate from an RCT [39], and ambiguous TIR unit scaling [40]. Study characteristics are in Table 1 [22–26,41–46]. Studies spanned 2020–2026 across five countries (China 4, Japan 3, South Korea 2, Belgium 1, Portugal 1). Seven were T2D, four T1D; ten cross-sectional, one retrospective cohort. Sample sizes ranged from 161 to 1,274 and CGM duration from 3 to 28 days. Four studies adjusted for HbA1c. ROBINS-E ratings were Some concerns (10) and High (1; Bezerra 2023). Full domain judgements are in Supplementary Table S1 (Supplementary Figures S2–S3). The pooled OR per 10% TIR was 0.8932 (95% KH CI 0.8420– 0.9476; P = 0.002; Figure 2), corresponding to approximately 11% lower odds of DKD per 10% TIR increment. Heterogeneity was substantial (I² = 69.6%; τ² = 0.00341; Q = 38.3, df = 10, P < 0.001) and the 95% prediction interval (0.77–1.03) crossed unity. The HbA1c-adjusted subgroup (k = 4) yielded OR 0.91 (95% CI 0.86–0.97; P = 0.015; I² = 0%); the association persisted with no detectable residual heterogeneity, indicating it is not fully attributable to average glycemia. This subgroup observation should be interpreted cautiously, as the formal test of HbA1c-adjustment status as a moderator was not significant (R² = 0%; P = 0.59) and I² is imprecisely estimated at k = 4. The association was significant in T2D (k = 7; OR 0.88; P = 0.002) and directionally protective in T1D (k = 4; OR 0.85; P = 0.25). Full subgroup results including geographic region, outcome type, and study design are in Table 2. Across all leave-one-out iterations, the pooled OR remained significant (range 0.88–0.91; all P ≤ 0.005), and excluding the single High-RoB study (Bezerra 2023) yielded a materially identical estimate (OR 0.90; 95% CI 0.84–0.95; P = 0.003). On influence diagnostics, two studies were flagged by the composite rule: Wakasugi 2021 (Cook’s distance 0.66) and Kim JY 2025 (DFFITS 0.66); both reflect high random-effects weight rather than outlier status, with studentized residuals within ±3 (Supplementary Table S3). The pooled estimate was robust across six alternative between-study variance estimators (OR range 0.88–0.90; all P < 0.003) and to replacing the KH adjustment with Wald inference. The Zhang HR-to-OR conversion, unconverted HR, and Zhang-excluded specifications all yielded significant and materially identical pooled estimates (OR 0.89, 0.90, and 0.89, respectively; Supplementary Table S3). No pre-specified moderator reached statistical significance in univariate meta-regression. Diabetes type was the only moderator with non-zero explained variance (R² = 33.3%; P = 0.29); all others had R² = 0%. Given the small number of studies relative to candidate covariates (k = 11), these exploratory analyses were insufficiently powered to detect all but very large moderation effects (Supplementary Table S4). Egger’s test detected significant funnel asymmetry (t = −4.89; df = 9; P = 0.001), whereas Begg’s rank correlation did not (τ = −0.31; P = 0.22). Duval-Tweedie trim-and-fill imputed four studies on the right side; the refit estimate remained statistically significant (OR 0.92, 95% CI 0.87–0.98; P = 0.006). The significant Egger’s test is noted in the GRADE assessment (Supplementary Figure S1). For the primary pool, certainty was Very Low (⊕○○○): downgraded for risk of bias (7/11 studies lacked HbA1c adjustment), inconsistency (I² = 69.6%; prediction interval crosses null), and publication bias (significant Egger’s test). For the HbA1c-adjusted subgroup, certainty was Low (⊕⊕○○), with one upgrade for plausible confounding direction. Domain-level rationales are in Table 3 and Supplementary Table S2.

**Fig. 2.**
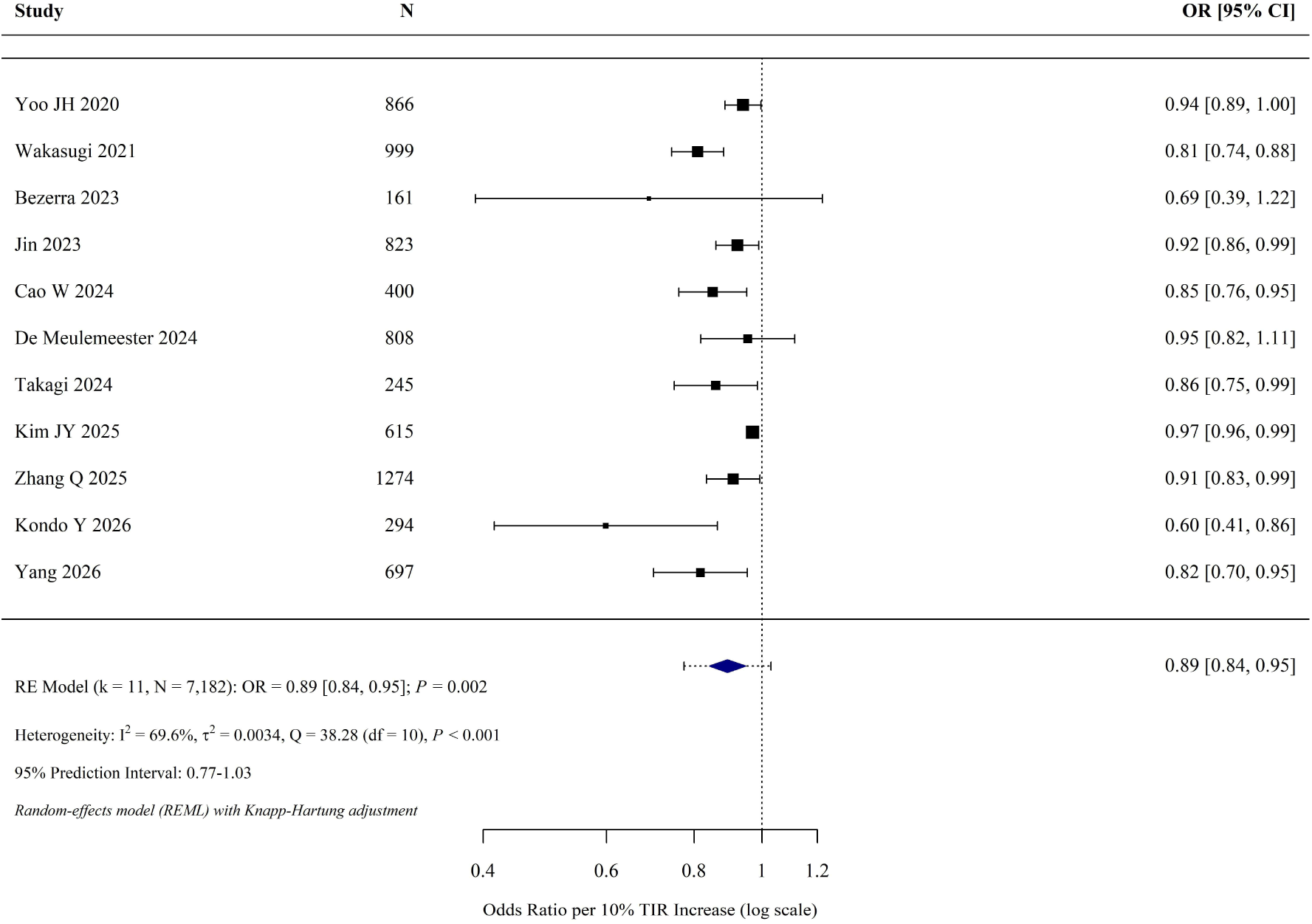
Forest plot of the association between CGM-derived time in range (per 10% increment) and diabetic kidney disease. Squares represent individual study odds ratios (size proportional to weight); horizontal lines indicate 95% confidence intervals. The diamond represents the pooled estimate from the random-effects model (REML with Knapp–Hartung adjustment). The dashed line indicates the prediction interval (0.77–1.03).

**Table 1.**
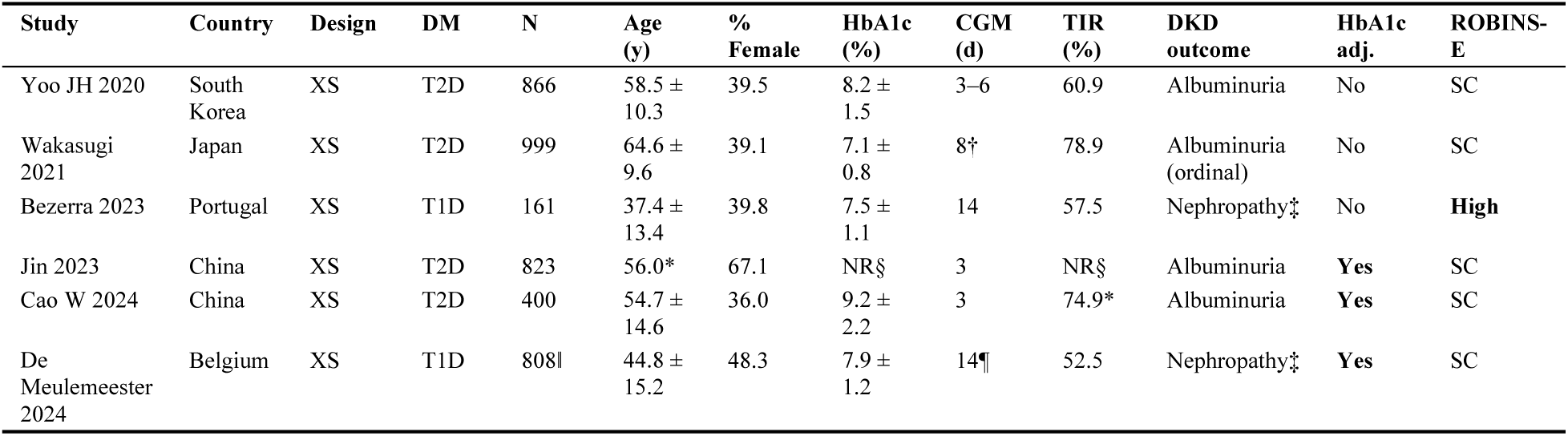
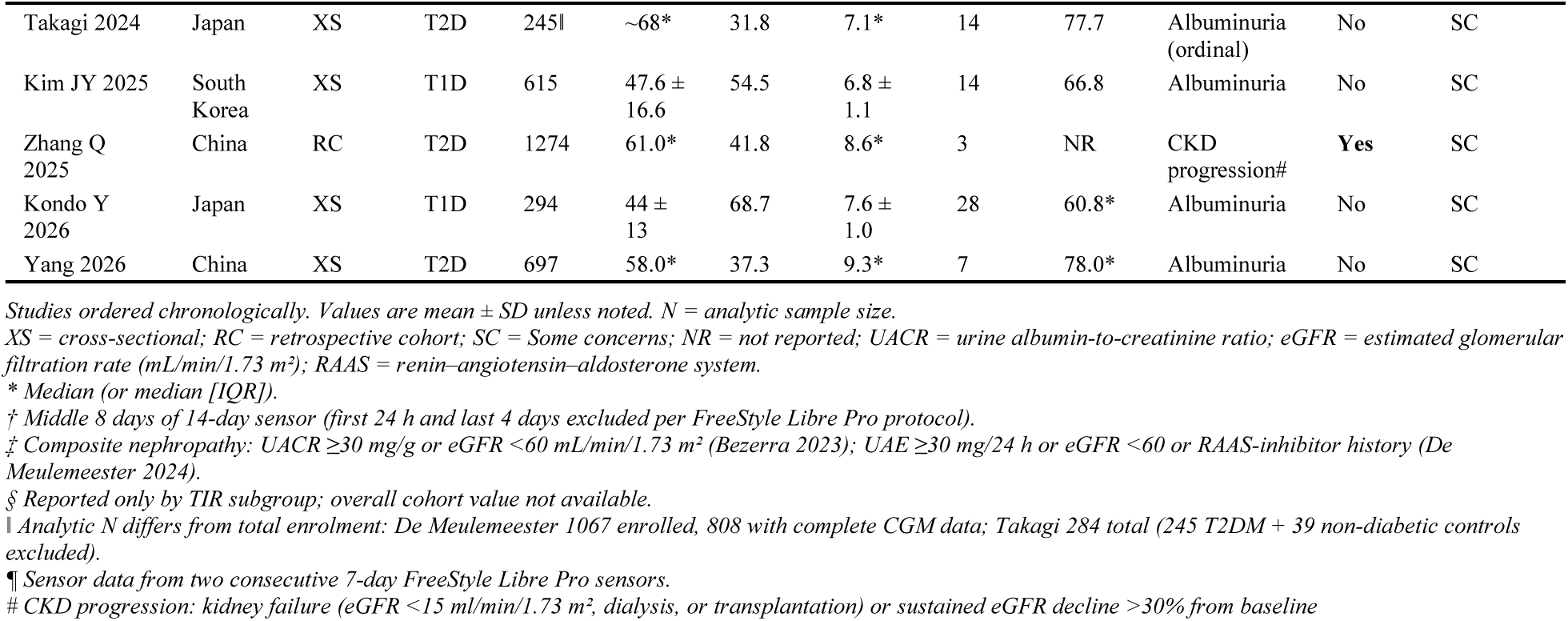
Characteristics of the 11 included studies.

**Table 2.** Subgroup analyses of the association between TIR (per 10% increase) and DKD.

| Subgroup | k | N | OR (95% CI) | P | I<sup>2</sup> (%) | P interaction |
| --- | --- | --- | --- | --- | --- | --- |
| **Overall (primary pool)** | **11** | **7,182** | **0.89 (0.84–0.95)** | **0.002** | **69.6** | **-** |
| *Diabetes type* |  |  |  |  |  |  |
| Type 2 diabetes | 7 | 5,304 | 0.88 (0.83–0.93) | 0.002 | 48.5 |  |
| Type 1 diabetes | 4 | 1,878 | 0.85 (0.59–1.22) | 0.25 | 80.8 | 0.29 |
| *HbA1c adjustment* |  |  |  |  |  |  |
| HbA1c-adjusted | 4 | 3,305 | 0.91 (0.86–0.97) | 0.015 | 0.0 |  |
| HbA1c-unadjusted | 7 | 3,877 | 0.87 (0.77–0.97) | 0.022 | 83.2 | 0.59 |
| *Geographic region* |  |  |  |  |  |  |
| Asian | 9 | 6,213 | 0.89 (0.83–0.95) | 0.004 | 75.2 |  |
| Non-Asian | 2 | 969 | 0.92 (0.23–3.65) | 0.57 | 14.0 | 0.73 |
| *Outcome definition* |  |  |  |  |  |  |
| Albuminuria | 8 | 4,939 | 0.88 (0.81–0.96) | 0.009 | 79.2 |  |
| Non-albuminuria* | 3 | 2,243 | 0.92 (0.80–1.04) | 0.10 | 0.0 | 0.66 |
| *Study design* |  |  |  |  |  |  |
| Cross-sectional | 10 | 5,908 | 0.89 (0.83–0.95) | 0.004 | 73.5 |  |
| Cohort | 1 | 1,274 | -† | - | - | - |

Random-effects models (REML) with Knapp–Hartung adjustment. *P* interaction from mixed-effects meta-regression with the subgroup variable as moderator.
\* Non-albuminuria outcomes: composite nephropathy (Bezerra 2023, De Meulemeester 2024) and CKD progression (Zhang Q 2025).
† Only one cohort study (Zhang Q 2025); not separately pooled.
OR = odds ratio per 10% TIR increase; CI = confidence interval; *I*<sup>2</sup> = between-study heterogeneity; TIR = time in range (70–180 mg/dL); DKD = diabetic kidney disease; REML = restricted maximum likelihood.

**Table 3.**
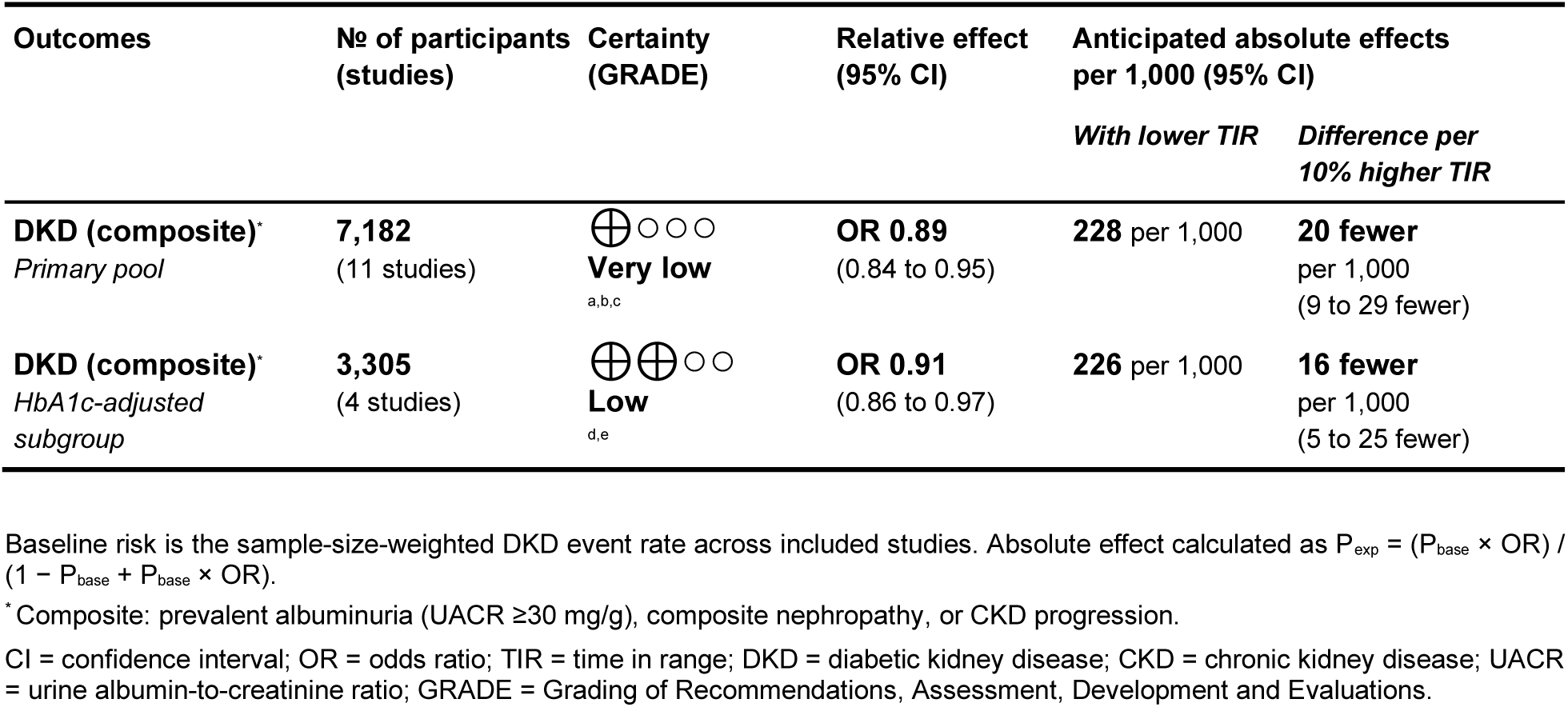
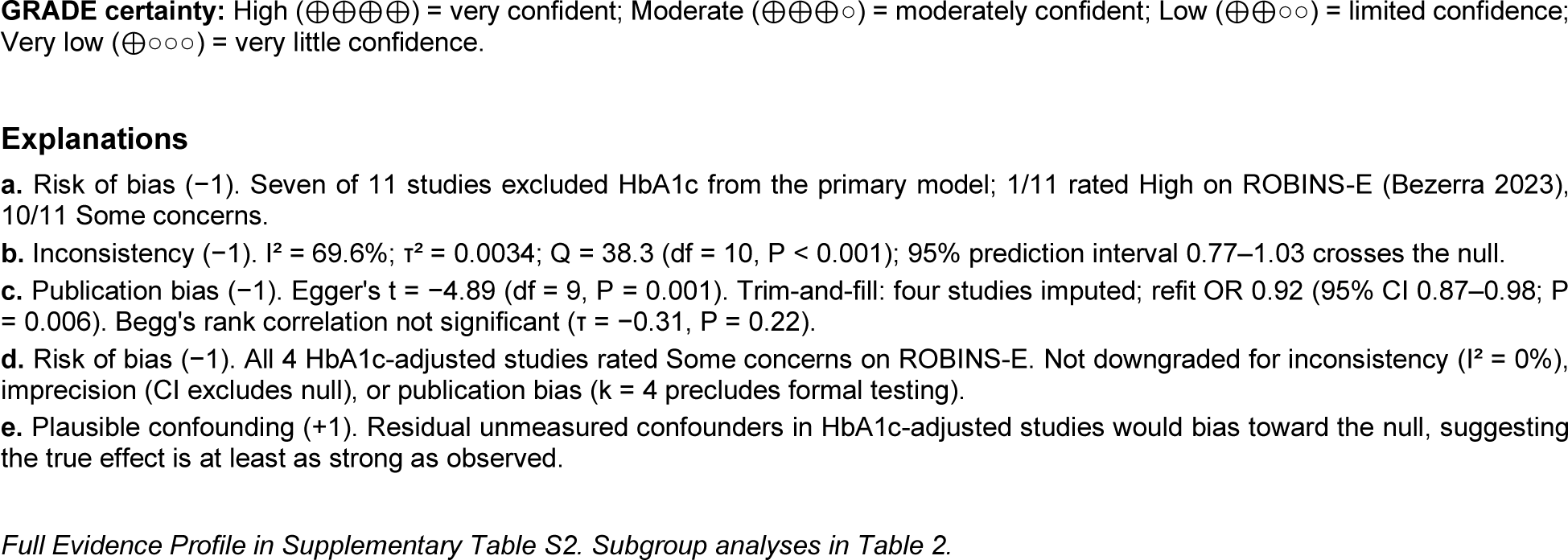
Summary of findings (GRADE)

## DISCUSSION

This meta-analysis pooled eleven observational studies including 7,182 adults with diabetes to estimate the association between CGM-derived time in range (TIR) and diabetic kidney disease (DKD). Each 10% higher TIR was associated with approximately 11% lower odds of DKD (OR 0.89, 95% CI 0.84–0.95). The direction of effect was consistent in every leave-one-out iteration and across seven between-study variance estimators. In the four studies that adjusted for HbA1c, the association remained statistically significant (OR 0.91, 95% CI 0.86–0.97; P = 0.015), and between-study heterogeneity fell to zero (I² = 0%). Under GRADE, certainty of evidence was Very Low for the full pool and Low for the HbA1c-adjusted subgroup. The substantial heterogeneity in the full pool (I² = 69.6%) reflects real differences between studies rather than random variation. Included studies varied in kidney-outcome definition, CGM monitoring duration (3 to 28 days), diabetes type, geographic setting, and whether the primary model adjusted for HbA1c. Despite this variation, all eleven studies reported point estimates below the null. The fact that heterogeneity disappeared once HbA1c was held constant suggests that confounding by average glycemia is a major source of between-study variability. No other moderator examined in meta-regression accounted for a comparable share of the variance. Earlier reviews of CGM and microvascular outcomes have not focused specifically on the kidney. Yapanis et al. provided a narrative review of TIR across diabetes complications [27], and Raj et al. summarized TIR as a predictor of microvascular complications in type 2 diabetes without pooling estimates [47]. Individual studies have linked TIR to cardiovascular mortality [16], carotid intima-media thickness [15], retinopathy [18], and peripheral nerve dysfunction [17]. Five included studies but not in the meta-analysis examined CGM-derived TIR in relation to kidney outcomes but were synthesized narratively owing to incompatible effect measures. In a one-year randomized trial of sensor-augmented pump therapy in 26 adults with T1D and albuminuria, each 10% TIR increase was associated with a 19% UACR reduction (P = 0.04), though only unadjusted coefficients were reported [36]. A cross-sectional study of hospitalized T2D patients using flash CGM found lower TIR categories associated with higher diabetic nephropathy prevalence but used non-standard TIR groupings that precluded per-unit effect extraction [37]. Among 5,901 T2D patients profiled by CGM-derived glucose patterns, the lowest-TIR cluster had the highest prevalence of DKD after adjustment for HbA1c [38]. A randomized trial of sitagliptin add-on therapy in 46 T1D adolescents with nephropathy observed parallel improvements in TIR and UACR over three months but did not report a direct per-unit TIR–kidney association [39]. A retrospective analysis of 422 T2D patients reported a strong protective TIR–DKD association (OR 0.18; 95% CI 0.05–0.64) but ambiguous TIR unit scaling precluded back-transformation to a standardized per-10% effect [40]. All five studies reported findings in the protective direction. The present review adds to the literature in two ways: a first pooled kidney-specific estimate, separation of TIR from mean glycemia through an HbA1c-adjusted subgroup. Several mechanisms support the biological plausibility of this association. Glucose oscillation produces greater endothelial dysfunction and oxidative stress than mean-matched sustained hyperglycemia [20], and glucose excursions accelerate the formation of advanced glycation end-products implicated in podocyte injury and glomerular basement membrane thickening. HbA1c reflects average glycemic exposure over several months and does not capture these short-term dynamics, a limitation Beck et al. described as the “fallacy of average” [8]. Contemporary reviews argue that postprandial excursions, variability, and hypoglycemia carry mechanistic weight that HbA1c cannot represent [10]. TIR captures both magnitude and excursion frequency and has been validated as a surrogate for microvascular risk [14]. A kidney-specific TIR signal is therefore consistent with this broader evidence. The clinical implications of the present findings are modest. International consensus statements endorse TIR with a target of ≥70% for most non-pregnant adults with diabetes [11,12], and the ADA Standards of Care include TIR as a complementary glycemic metric [7]. The present findings support consideration of TIR in kidney-related risk stratification, particularly in patients whose HbA1c is acceptable, but whose glycemic variability may remain high. They do not, however, justify any change to HbA1c-based KDIGO 2022 algorithms [4]. TIR should be regarded as a useful adjunct to HbA1c in the assessment of kidney risk rather than as an independent treatment target. The strengths of the review include a pre-specified protocol, random-effects meta-analysis with REML and the Knapp–Hartung adjustment [33], which is preferred to DerSimonian–Laird in the presence of heterogeneity, and an extensive set of sensitivity, influence, and publication-bias analyses. Several limitations should be acknowledged. All eleven studies were observational; only one used a retrospective cohort design with longitudinal follow-up [25]; the remainder were cross-sectional. None used hard kidney endpoints such as end-stage renal disease or renal death. Only five of the eleven studies met the international consensus recommendation of ≥14 days of CGM monitoring for reliable TIR estimation; shorter monitoring biases the estimate toward the null, which suggests that the true effect is at least as large as the pooled value. Only four studies adjusted for HbA1c, which limits both the precision and the generalizability of the key subgroup analysis. Nine of the eleven cohorts were Asian, which restricts generalizability to other populations. Funnel asymmetry was significant on Egger’s test (P = 0.001), although Begg’s rank correlation was not (P = 0.22) and trim-and-fill imputed four studies, and the refit estimate remained significant (OR 0.92; P = 0.006), leaving the pooled estimate unchanged. Meta-regression across eleven studies is under-powered, and the moderator analyses should therefore be regarded as hypothesis-generating. The 95% prediction interval (0.77 to 1.03) crosses unity, indicating that a future study of similar design could plausibly observe a null association. Future research should address these limitations directly. Prospective cohort studies should adopt at least 14 days of CGM monitoring, HbA1c-adjusted primary models, and KDIGO-compliant outcome adjudication including hard endpoints such as end-stage renal disease and sustained eGFR decline [4]. A randomized trial comparing TIR-targeted glycemic management (≥70% per international consensus) with HbA1c-targeted usual care, with pre-specified kidney endpoints, would be required to establish causality. The 2023 international consensus statement endorses CGM-derived metrics, including TIR, as primary or key secondary endpoints in clinical trials [48], providing a methodological framework for such a study. An individual-patient-data meta-analysis of the studies synthesized here would allow harmonized outcome definitions, modeling of dose–response relationships, and identification of patient-level effect modifiers. In summary, this meta-analysis identifies a consistent inverse association between CGM-derived TIR and DKD across diverse observational studies. The persistence of the association after adjustment for HbA1c, together with the resolution of heterogeneity in this subgroup, indicates that TIR carries information about kidney risk that is not fully captured by HbA1c. The strength of the evidence remains limited, and randomized trials with hard kidney endpoints will be needed before TIR can be considered a target for kidney-protective glycemic management.

## CONCLUSIONS

In this first quantitative meta-analysis of CGM-derived TIR and DKD, each 10% increase in TIR was associated with approximately 11% lower odds of DKD (OR 0.89, 95% CI 0.84–0.95; P = 0.002) across 11 observational studies (N = 7,182). The four HbA1c-adjusted studies showed a statistically significant association with no detectable residual heterogeneity (OR 0.91, 95% CI 0.86–0.97; P = 0.015; I² = 0%), indicating the signal is not fully explained by average glycemia, while acknowledging that this subgroup is small and the corresponding moderator test was not significant. No pre-specified moderator explained heterogeneity. The finding was robust to outlier exclusion and design restriction; trim-and-fill imputed four studies (refit OR 0.92, 95% CI 0.87– 0.98; P = 0.006) (OR unchanged). GRADE certainty was Very Low for the primary pool and Low for the HbA1c-adjusted subgroup. Prospective cohorts with standardized CGM protocols, HbA1c-adjusted primary analyses, and KDIGO-compliant outcome adjudication are needed to move from association to causation.

## Abbreviations

ADA: American Diabetes Association
AGE: advanced glycation end-product
CGM: continuous glucose monitoring
CI: confidence interval
CKD: chronic kidney disease
CV: coefficient of variation
DCCT: Diabetes Control and Complications Trial
DKD: diabetic kidney disease
eGFR: estimated glomerular filtration rate
GRADE: Grading of Recommendations, Assessment, Development and Evaluations
HbA1c: hemoglobin A1c
HR: hazard ratio
I²: inconsistency statistic
KDIGO: Kidney Disease: Improving Global Outcomes
KH: Knapp–Hartung
OR: odds ratio
PECO: Population, Exposure, Comparator, Outcome
PRISMA: Preferred Reporting Items for Systematic Reviews and Meta-Analyses
PROSPERO: International Prospective Register of Systematic Reviews
Q: Cochran’s Q statistic
RAAS: renin–angiotensin–aldosterone system
RC: retrospective cohort
REML: restricted maximum likelihood
ROBINS-E: Risk of Bias in Non-randomized Studies of Exposures
RR: risk ratio
SC: Some concerns
SE: standard error
SMBG: self-monitoring of blood glucose
T1D: type 1 diabetes
T2D: type 2 diabetes
TIR: time in range
UACR: urine albumin-to-creatinine ratio
UKPDS: United Kingdom Prospective Diabetes Study
XS: cross-sectional
τ²: between-study variance.

## Acknowledgments

None.

## CRediT author statement

SML: Conceptualization, Methodology, Software, Formal analysis, Investigation, Data curation, Writing-original draft, Visualization. SK: Investigation, Data curation, Validation, Writing-review & editing. SA: Investigation, Validation, Writing-review & editing. PD: Investigation, Validation, Writing-review & editing. SG-Conceptualization, Validation, Writing-review & editing.

## Data availability

All data analyzed in this study were extracted from published articles. The extracted dataset, R analysis scripts, and data collection forms are available from the corresponding author upon reasonable request.

## Ethics approval

Not Applicable

## Consent to Participate

Not Applicable

## Consent for publication

Not Applicable

